# Favipiravir for treating novel coronavirus (COVID-19) patients: protocol for a systematic review and meta-analysis of controlled trials

**DOI:** 10.1101/2020.04.27.20081471

**Authors:** Morteza Arab-zozani, Soheil Hassanipour, Djavad Ghoddoosi-Nejad

**Affiliations:** Social Determinants of Health Research Center, Birjand University of Medical sciences, Birjan, Iran.; Gastrointestinal and Liver Diseases Research Center, Guilan University of Medical Sciences, Rasht, Iran.; Department of Public Health, School of Health, Birjand University of Medical sciences, Birjan, Iran.

## Abstract

**Introduction:** An outbreak of severe acute respiratory syndrome coronavirus-2 (SARS-CoV-2) was reported in Wuhan, China in mid-December 2019, and declared a pandemic by the World Health Organization (WHO) on March 11, 2020. Due to the unknown nature of the disease and the lack of specific drugs, several potential treatments were used for patients. This systematic review and meta-analysis will evaluate studies of the effects of Favipiravir in COVID-19 pneumonia.

**Methods and analysis:** We will search electronic databases including LitCovid hub, PubMed, Scopus, ISI web of Sciences, Cochrane, and Embase using keywords related to COVID-19 and Favipiravir. We will search the reference lists of all included studies and reviews. We will also search for clinical trial registries, such as clinicaltrial.gov for the ongoing clinical trials. Two investigators (MAZ and SH) will independently screen titles, abstracts, and full-text of included studies based on eligibility criteria. These investigators will also independently extract data and appraise the quality of studies. All potential discrepancies will be resolved through consultation with the third reviewer. Data synthesis will be conducted using the Review Manager software (version 5.3) or CMA (version 2). Statistical heterogeneity will be assessed using a standard I^2^ test. A funnel plot, Egger’s test, and Begg’s test will be used for asymmetry to explore possible publication bias.

**Ethics and dissemination:** The findings of this systematic review with proportional meta-analysis will help to identify the safety and efficacy of Favipiravir for COVID-19 patients. Knowledge gained from this research will also assist physicians in selecting better treatment options and developing a guideline in this field.

**Strengths and limitations of this study:**

- In the protocol, all stage of study conducted by two reviewers independently and supervised by a third reviewer.
- This systematic review may produce the first meta-analysis that provides evidence regarding the safety and effectiveness of favipiravir on COVID-19 patients.
- The small number of studies published in this field when writing a protocol can be one of the most important limitations.

PROSPERO registration number: CRD42020180032

## Introduction

An outbreak of severe acute respiratory syndrome coronavirus-2 (SARS-CoV-2) was reported in Wuhan, China in mid-December 2019, and declared a pandemic by the World Health Organization (WHO) on March 11, 2020^1, 2^. Novel coronavirus 2019, named COVID-19 by WHO, characterized by several symptoms including fever, cough, and shortness of breath. This disease is more severe in men, the elderly, and people with other chronic health conditions, such as high cardiovascular disease, diabetes, chronic respiratory disease, and hypertension ^3, 4^. As of 14 April 2020, nearly 2,000,000 people have been diagnosed with COVID-19 and about 120,000 deaths in the world ^5^.

As escalating pandemic of COVID-19 and potential impact on global health preparing effective therapeutic options is urgently needed^6^. In addition to other drugs such as lopinavir, ritonavir, ribavirin, and chloroquine phosphate, which are used to treat this disease, the use of Favirapivir is also being initiated in many clinical trials ^7, 8^. Favipiravir is a purine nucleic acid analog and virus RNA dependent RNA polymerase (RdRp) inhibitors that sold under the brand name Avigan, which is an antiviral medication approved in 2014 by Japan Pharmaceuticals and Medical Devices Agency for the treatment of influenza A virus infection. It is also being studied to treat several other viral infections including COVID-19^9, 10^.

While the use of Favipiravir drug is being applied for the treatment of COVID-19 patients, uncertainty remains about its safety and effectiveness. Therefore, we aim to systematically review the available literature of the application of Favipiravir in COVID-19 patients to examine the empirical evidence of the effects of this drug for COVID-19 pneumonia. We intend to provide vigorous evidence for clinical practice in treating COVID-19 patients.

## Methods and analysis

### Protocol and registration

This protocol has been registered in the PROSPERO International Prospective Register of Systematic Reviews (CRD42020180032), on 5 February 2020. We have arranged this protocol following the Preferred Reporting Item for Systematic Review and Meta-analysis (PRISMA-P) statement (Appendix 1) ^11^. Also, this systematic review and meta-analysis will report according to the Preferred Reporting Items for Systematic Reviews and Meta-Analyses (PRISMA) statement ^12^.

### Eligibility criteria

All types of clinical trials (study design) that have investigated the safety and efficacy of Favipiravir (intervention) compared with other control groups (comparison) for treatment of patients with confirmed infection with SARS-CoV2 (population) will be included. There will be no restrictions concerning gender, age, ethnicity, blinding, follow-up, or publication status. Publications in English and Farsi will be included. Survival of the patients at the end of treatment and follow up will be the primary outcome, followed by the time and rate of the patient with a negative test for the COVID-19. Additional outcomes will consist of a decreased rate of symptoms, proportion transferred to the ICU, length of stay in the hospital, ICU length of stay, quality of life, and adverse events (outcomes). Articles with unavailable full text in English or Farsi languages or whose full text is not accessible will be excluded from the study. Also, studies that have insufficient or incomplete data will not be incorporated.

### Information sources and search strategy

Two independent reviewers (MA-Z and SH) will search electronic databases including PubMed, Scopus, ISI web of Sciences, Cochrane, Embase, LitCovid hub ^13^, and Scientific Information Database^14^ using keywords combination (MeSH term and free term), such as “2019 nCoV” OR 2019nCoV OR “2019 novel coronavirus” OR COVID-19 OR “new coronavirus” OR “novel coronavirus” OR “SARS CoV-2” OR (Wuhan AND coronavirus) OR “SARS-CoV” OR “2019-nCoV” OR “SARS-CoV-2” and Favipiravir OR Avigan. We will search the reference lists of all included studies, reviews, and clinical trial registries, for an ongoing clinical trial (see Appendix 2 for the final proposed PubMed search strategy).

### Study records

After importing records to EndNote X7 software and removing duplicate records, two reviewers (SH and DGh) will independently screen titles, abstracts and full-texts of included studies based on predefined eligibility criteria to identify studies concerning safety and efficacy of Favipiravir among patients with COVID-19. A kappa (κ) statistic will be used to calculate the extent of inter-observer agreement on the independent inclusion of articles. All potentials discrepancies will be resolved by consultation with a third reviewer (MA-Z).

### Data extraction and data items

Two reviewers (SH and DGh) will independently extract data from included studies using a pre-piloted data extraction form. We will pilot this form using at least three examples of included studies, and if there is an agreement above 90%, it will be approved. The data extraction form includes the following items; authors name, year of the publication, study design, study sample, country of origin, mean age of participants, gender, the severity of diseases, comorbidities, type of intervention and dose, control group, follow up, randomization, blinding, allocation concealment, primary and secondary outcomes, and adverse events. All potentials discrepancies will be resolved by consultation with a third reviewer (MA-Z).

### Risk of bias in individual studies

Two reviewers (SH and DGh) will independently assess the risk of bias among the included studies. We will assess the risk of bias of the included studies using Cochrane Collaboration criteria including seven items of selection bias (random sequence generation and allocation concealment), performance bias, detection bias, attrition bias, reporting bias, and other bias. Any discrepancies will be resolved by consultation with a third reviewer (MA-Z).

### Data synthesis

Statistical analyses will be carried out using the RevMan software (version 5.3) or CMA (version 2). We will conduct analyses employing risk ratios (RR) and mean differences (MD) with 95% confidence intervals for dichotomous continuous data, respectively. Statistical heterogeneity will be tested using Cochran’s Q statistic and quantified using the I^2^ statistic. If possible, we will perform subgroup analyses based on dose, follow up time, level of disease and age group. The Mantel-Haenszel method and the DerSimonian and Laird inverse variance method will be used for dichotomous outcomes and continuous outcomes, respectively. The fix or random-effects model will be used to pool the data based on the level of heterogeneity and the number of studies in each unit of analyses. A funnel plot, Egger’s test, and Begg’s test will be used for asymmetry to explore possible publication bias.

## Patients and public involvement

We will not collect primary data, then, ethical approval will not be required.

## Ethics and dissemination

The onset of COVID-19 and its subsequent pandemic situation is becoming a substantial global health emergency ^15^. This systematic review and meta-analysis will be carried out to investigate the world’s relevant literature on the safety and effectiveness of Favipiravir in the treatment of COVID-19 patients. Favipiravir, a purine nucleic acid analog, and potent RdRp inhibitor played an important role in the treatment of influenza and Ebola in recent years ^9^. Several drugs such as chloroquine, arbidol, remdesivir, and favipiravir are currently undergoing clinical studies to test their efficacy and safety in the treatment of coronavirus disease 2019 in many countries such as Iran, Japan and China ^7, 8^. To date, there is no gold standard for the treatment of COVID-19 as evidence is poor ^16^. The findings of this systematic review and meta-analysis will help to evaluate the potential safety and effectiveness of Favipiravir compared to other drugs. We hope the knowledge gained from this research will also assist physicians in selecting better treatment options and developing a guideline in this field.

## Data Availability

All related data exist in protocol. The complete data will be published after completing the review

## Patient consent for publication

Not applicable

## Competing interests

The authors declare that they have no competing interests.

## Acknowledgements

None

## Funding

None

## Contributors

MA-Z designed the review. MA-Z and SH conducted the database searches. SH and DGh screened the included studies, Pilot data extraction form, extracted data, and quality appraisal. MA-Z and SH performed meta-analysis. MA-Z and SH wrote the manuscript draft. All authors read, revised and approved the final manuscript.

## Notes

### Competing Interest Statement

The authors have declared no competing interest.

